# Characteristics and Outcome of Immunoglobulin A Nephropathy – a Swiss single center experience

**DOI:** 10.1101/2025.06.04.25328997

**Authors:** Danny T. Taing, Bruno Vogt, Laila-Yasmin Mani

## Abstract

**Background:** Immunoglobulin A nephropathy (IgAN) is the most common primary glomerulonephritis worldwide. Geographic differences in disease course and treatment response are well recognized. The purpose of this analysis was to study clinical and histological characteristics, treatment practices and outcome of IgAN cases from a Swiss tertiary center.

**Methods:** This retrospective cohort analysis identified 158 cases of adult biopsy-proven IgAN by chart review diagnosed between 1980 and 2017. Following detailed phenotypisation, standard descriptive methods and univariate analysis corrected by the Bonferroni method were applied.

**Results:** Patients were majorly male and of Caucasian descent. At diagnosis, mean estimated glomerular filtration rate (eGFR) was 55.4 ml/min/1.73 m^2^, mean proteinuria was 2.4 g/d, 69.9% of the patients were hypertensive. Clinical presentation varied according to age. Initial biopsies showed moderate to severe tubular atrophy and interstitial fibrosis (IFTA) in 29.1% and crescents in 36.7% of cases. Therapy included renin-angiotensin-aldosterone-inhibitors in 86.7%, immunosuppressive therapy in 46.8% including steroids and other immunosuppressive drugs (28.7%), mainly azathioprin. Outcome included 34.1% complete and 22.2% partial remissions, relapses in 32.0% of patients, while 43.0% of patients progressed to ESKD during follow-up (median 100.0 months). Recurrence rate after transplantation was 18.8%. Immunosuppressive therapy was more frequently used in patients with higher proteinuria level and crescents. Predictors of progression were lower eGFR, higher proteinuria and higher extent of IFTA on the initial biopsy.

**Conclusions:** This retrospective cohort analysis gives insight into characteristics and outcome of patients with IgAN from a Swiss tertiary center, treatment practices as well as predictors of outcome and therapy choices.

**Key learning points:** *What was known:*

- A wide range of clinical presentations exists for immunoglobulin A nephropathy (IgAN) pointing to IgAN as a disease spectrum rather than a single disease entity
- Additional important geographic differences regarding disease prevalence, disease course and response to therapies have been recognized
- Progression to end-stage kidney disease (ESKD) occurs in a substantial part of affected patients

*This study adds:*

- In this cohort, a comparatively high proportion of patients was treated by immunosuppressive therapy including non-steroid treatment underlining the heterogeneous nature of treatment practices worldwide
- Predictors of the use of immunosuppressive therapy in this cohort have been identified
- Similarly to recent UK registry data, a high rate of progression to ESKD has been found including patients with low baseline proteinuria level

*Potential impact:*

- These findings further underline the importance of achieving an improved characterization and pathogenetic understanding of IgAN disease subtypes through ongoing research efforts
- At the beginning of the current new therapeutic era for patients with IgAN, this goal has now become more relevant than ever

## Introduction

Immunoglobulin A (IgA) nephropathy is the most common primary glomerulonephritis worldwide [1, 2]. It is defined histologically by the dominant or co-dominant glomerular deposition of IgA1 antibodies in a mesangial location [3]. Progression to end-stage kidney disease (ESKD) occurs in a substantial part of affected patients [4]. However, the range of clinical manifestations is wide reaching from asymptomatic isolated urinary abnormalities, progressive chronic impairment of kidney function or nephrotic syndrome to rapid progressive acute kidney injury (AKI) and systemic vasculitis (Henoch-Schönlein purpura) [5, 6]. Moreover, large geographic and ethnic differences exist with regard to disease prevalence, presentation, outcome, and response to therapy [5, 7-12]. Taken together, these data suggest IgA nephropathy as a disease spectrum rather than a well-defined disease entity [13, 14].

The primary form of IgA nephropathy is an immune-mediated disease involving a multi-hit pathogenesis [15]. Until recently, treatment approaches focused on supportive care including renin-angiotensin-aldosterone-system (RAAS) inhibition, more recently sodium-glucose cotransporter-2 inhibition and optimal control of cardiovascular risk factors [16, 17]. The role of an autoimmunity-oriented approach as opposed to a purely chronic kidney disease (CKD)-oriented approach has been controversially debated during the last decades [18, 19]. Given the paucity of high quality clinical data and concern for medication-induced toxicity, immunosuppressive treatment had generally been reserved for patients with high risk of disease progression or rapid progressive disease course [20]. However, local therapy regimens greatly varied according to geographic location and probably even according to centers [21].

In this work, we aimed to perform a detailed review of all cases of patients with IgA nephropathy treated at a Swiss tertiary center with focus on disease characteristics, treatment practices and outcome as well as to evaluate potential predictors of therapeutic choices and patient outcome.

## Material and Methods

The study was designed as a retrospective cohort analysis including all adult patients with a diagnosis of biopsy-proven IgA nephropathy included in the electronic medical record (introduced in November 2008) of the University clinic for Nephrology and Hypertension at the University Hospital Bern as of December 7^th^ 2017. The electronic medical record database was searched using the term “IgA”. Inclusion criteria were patients with a diagnosis of IgA nephropathy older than 18 years old. Exclusion criteria were lack of performed kidney biopsy and lack of clinical data entries.

For eligible patients, chart review was performed by collecting relevant pre-defined disease-related data within the electronic medical records. The dataset contained the following parameters:

- demographic parameters (age, sex as noted in the clinical chart, ethnicity, place of birth)
- family history (e.g. kidney disease, consanguinity)
- clinical parameters (height, body weight, body mass index, blood pressure)
- clinical presentation (renal and extrarenal manifestations)
- laboratory parameters (hemoglobin, serum albumin, serum creatinine, estimated glomerular filtration rate (eGFR) using Chronic Kidney Disease Epidemiology Collaboration (CKD-EPI) formula, CKD stage according to Kidney Disease: Improving Global Outcomes, measured creatinine clearance, 24-hour urine protein, 24-hour urine albumin, protein-creatinine-ratio (g/mol), albumin-creatinine-ratio, hematuria intensity)
- histological parameters (proportion of glomeruli with crescents, Haas classification [22] stage and Oxford classification/MEST score [23], tubular atrophy and interstitial fibrosis (IFTA) grading, IgA/IgG and complement C3 deposit grading
- administered therapies (RAAS inhibitors, steroids, immunosuppressive drugs, tonsillectomy)
- outcome data (remission, ESKD, dialysis or kidney transplantation, recurrence of IgA nephropathy in the kidney transplant, death), as well as
- clinical and laboratory parameters at the time of last follow-up.

*Complete remission* was defined as sustained proteinuria < 0.3 g/d with preserved kidney function within one year after treatment start. *Partial remission* was defined as ≥ 50% decrease in proteinuria and proteinuria ≥0.3 g/d and <1 g/d within one year after treatment start. *No remission* was defined as not fulfilling criteria for remission. *Relapse* was defined as the reappearance of proteinuria >1g/d and increase of 50% from the lowest level of proteinuria in remission. *Acute kidney injury* was defined according to Kidney Disease Improving Global Outcomes Guidelines 2012 [24].

Statistical analysis was performed using Microsoft Excel® 2016 and included standard descriptive methods (means, standard deviation or medians and ranges as appropriate). Continuous variables were compared using student’s t-test on a two-sided significance level of 0.05. Dichotomic variables were compared using Chi square test. Correction for multiple testing was performed using the Bonferroni method.

The study was approved by the cantonal ethics committee of the canton of Bern, Switzerland (approval number #2022-02292).

## Results

### Patient cohort

The screening of the hospital’s electronic database for the term “IgA” between 1980 and 2017 resulted in 2543 patient entries. Among those, 2374 had no diagnosis of IgA nephropathy and were therefore excluded. Another 11 patients were excluded because of lack of histological confirmation or of missing clinical data. Finally, a total of 158 patients diagnosed between 1980 and 2017 was included in the analysis (Figure 1).

**Figure 1.**
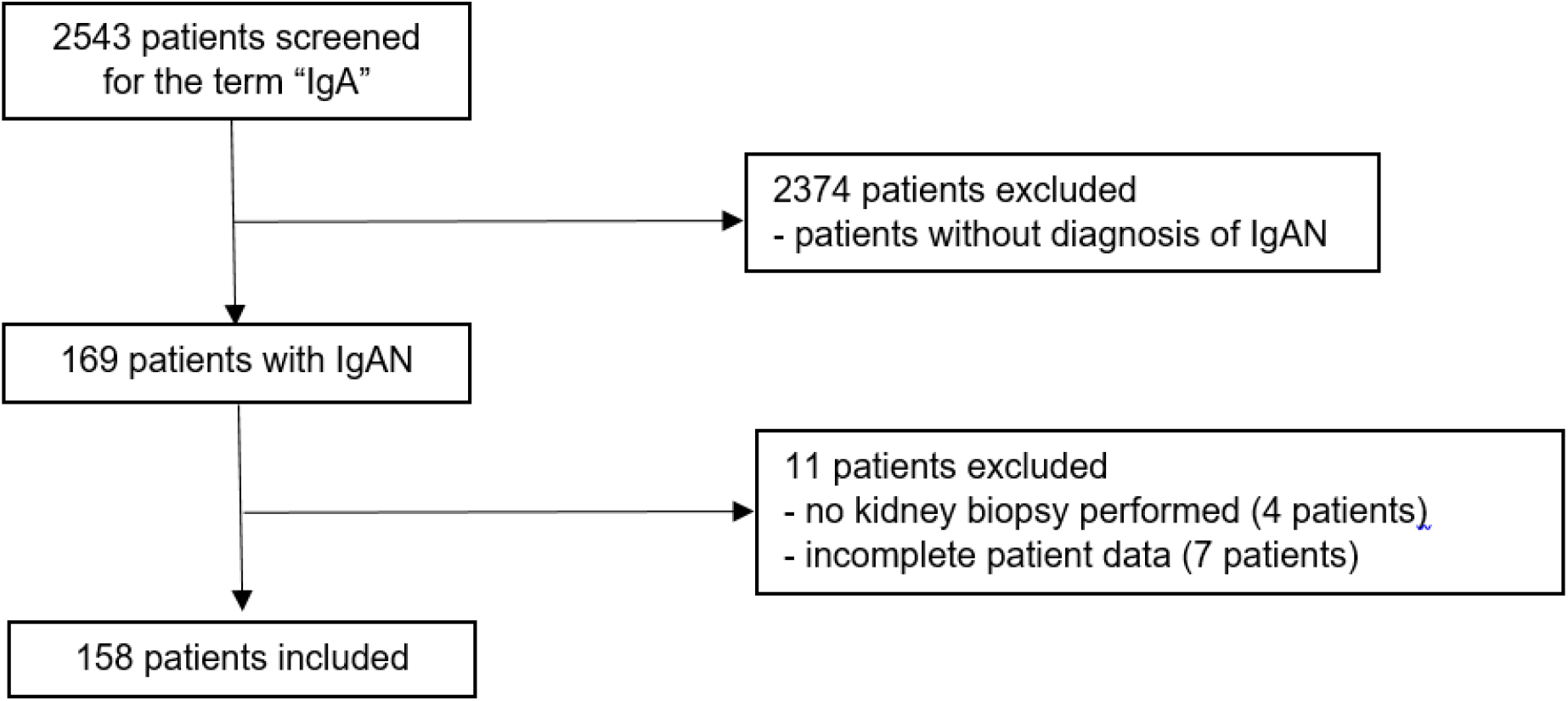
Patient flow chart. IgAN: IgA nephropathy.

### Baseline characteristics, clinical and histological presentation

Baseline characteristics of the included patients are shown in Table 1. Patients were predominantly male and of Caucasian ethnicity. Positive family history for IgA nephropathy was present in 0.6% of patients.

**Table 1:** Baseline patient characteristics at diagnosis (n=158)

| Demographics and basic clinical data |  |
| --- | --- |
| Age (years) | 53.5 ± 14.8 |
| Gender (%) |  |
| - female | 21.1 |
| - male | 78.9 |
| Ethnicity (%) |  |
| - Caucasian | 85.4 |
| - Asian | 8.9 |
| - African | 3.2 |
| - Hispanic | 2.5 |
| Positive family history for IgA nephropathy (%) | 0.6 |
| Clinical presentation |  |
| Blood pressure (mmHg) |  |
| - systolic (n=127) | 138.3 ± 21.9 |
| - diastolic (n=127) | 84.0 ± 14.2 |
| Hypertension (%) (n=109) | 69.9 |
| Serum creatinine (μmol/l) (n=136) | 167.2 ± 107.2 |
| Estimated glomerular filtration rate* (ml/min/1.73 m<sup>2</sup>) (n=136) | 56.8 ± 29.9 |
| Proteinuria<sup>+</sup> (g/d) (n=132) | 2.4 ± 2.0 |
| Nephrotic syndrome (%) | 9.5 |
| Presence of microhematuria (%) (n=125) | 90.4 |
| Presence of macrohematuria (%) (n=153) | 29.1 |
| Extrarenal symptoms (%) (n=9) | 0.06 |
| Histological characteristics |  |
| Presence of crescents (%) (n=151) | 36.7 |
| Presence of moderate to severe tubular atrophy and interstitial fibrosis (%) (n=141) | 29.1 |

Values are expressed as means with standard deviation or proportions. Extrarenal symptoms included purpura, arthritis and abdominal pain. \*according to Chronic Kidney Disease Epidemiology Collaboration formula. <sup>+</sup>measured from adequately collected 24h-urine collection if available or estimated from urinary protein/creatinine ratio.

At presentation, nearly one third of patients was symptomatic including macrohematuria (29.1%), edema (19.0%) or flank pain (9.5%). Roughly 4% of patients presented with extrarenal symptoms including purpura and abdominal pain. The majority (69.9%) of patients were hypertensive. As expected, hematuria was seen in almost all patients, while over half of the patients presented with proteinuria ≥ 1 g/d with an overall mean proteinuria of 2.4 g/d. Almost one of ten patients had nephrotic syndrome at the initial presentation. Baseline kidney function ranged from preserved to severely impaired with a mean eGFR according to CKD-EPI across the cohort of 56.8 ml/min/1.73 m^2^. Three patients required dialysis treatment at presentation. Overall, most of the patients (86.7%) presented with chronically impaired kidney function.

Clinical presentation varied according to age. Thus, younger patients (18-39 years) presented significantly more frequently with macrohematuria and asymptomatic urine abnormalities and by trend acute kidney injury at time of diagnosis while nephrotic syndrome and extrarenal symptoms were recorded in similar proportions of patients (Figure 2a).

**Figure 2a.**
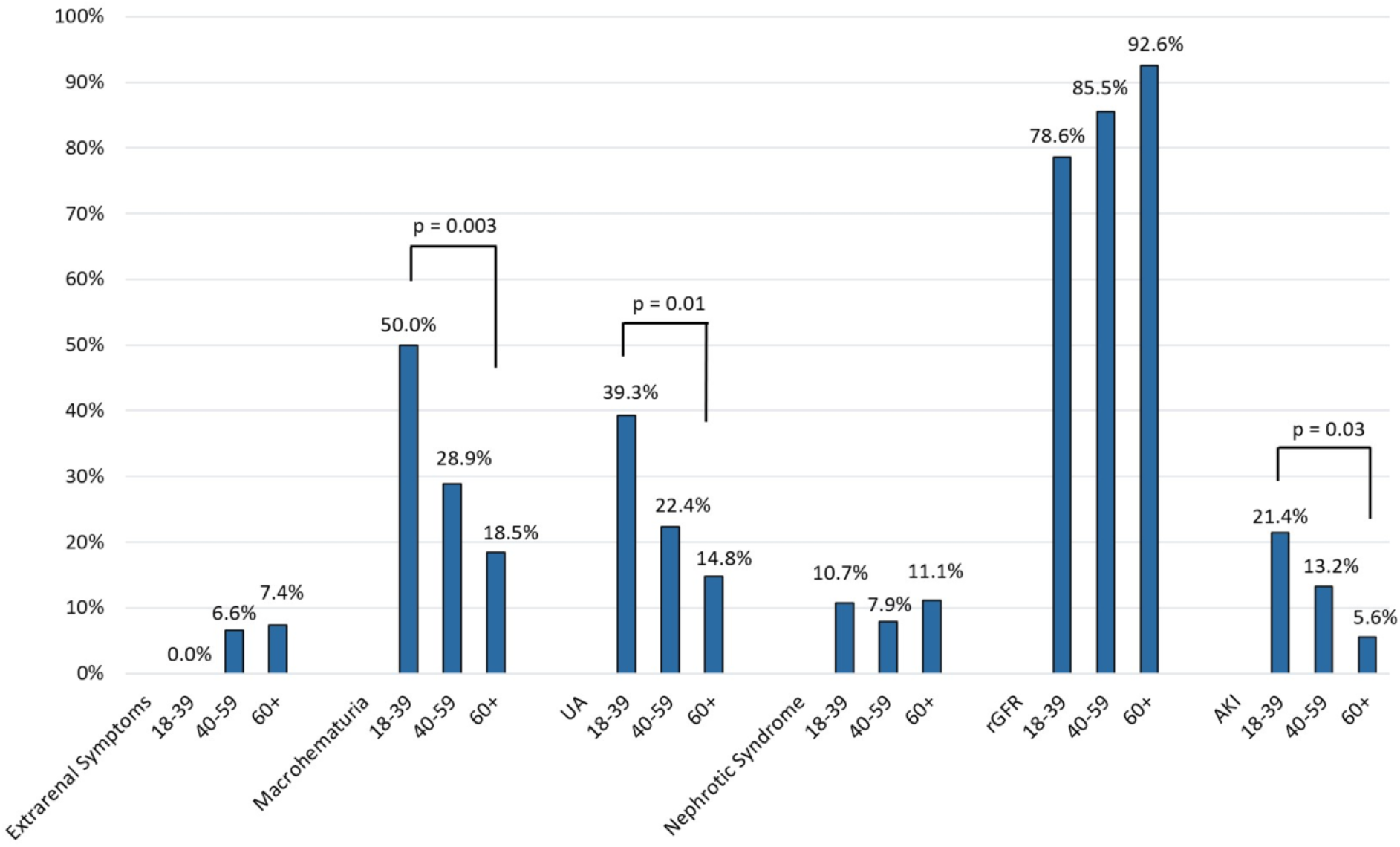
Clinical presentation according to age. Values are expressed as percentage of patients in each age group presenting with a particular clinical picture (mutually exclusive). Age groups: 18-39 years (n = 28), 40-59 years (n = 76), 60+ years (n = 54). Extrarenal symptoms included purpura, arthritis and abdominal pain. UA: Asymptomatic urine abnormalities. rGFR: reduced glomerular filtration rate (<60 ml/min/1.73 m^2^). Age group comparisons were performed using the Chi square-test without correction (if not indicated, p-values>0.05 were observed).

Furthermore, clinical presentation varied according to sex. By trend, women presented more frequently with nephrotic syndrome and asymptomatic urine abnormalities whereas macrohematuria tended to be the more likely presentation in men (Figure 2b).

**Figure 2b.**
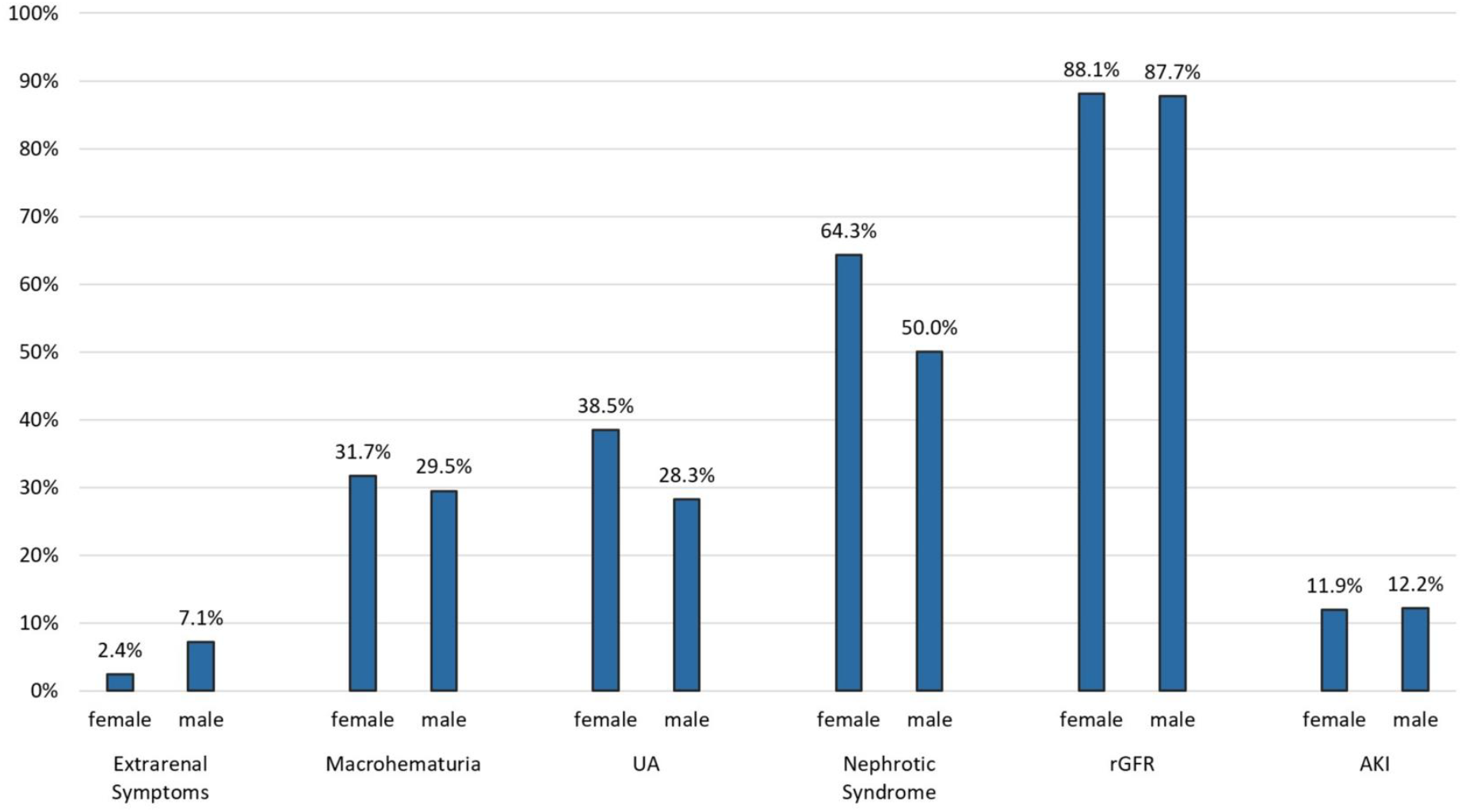
Clinical presentation according to sex. Values are expressed as percentage of patients in each age group presenting with a particular clinical picture (mutually exclusive). Age groups: 18-39 years (n = 28), 40-59 years (n = 76), 60+ years (n = 54). Extrarenal symptoms included purpura, arthritis and abdominal pain. UA: Asymptomatic urine abnormalities. rGFR: reduced glomerular filtration rate (<60 ml/min/1.73 m^2^). Group comparisons were performed using the Chi square test yielding p-values > 0.05.

The majority of kidney biopsies was analyzed before introduction of the Oxford classification. In 36.7% of biopsies, crescents were present; in 1.9% of biopsies, more than 50% of the glomeruli were affected by crescents. Moderate to severe tubular atrophy and interstitial fibrosis was noted in roughly a third of patients on the initial biopsy.

### Treatment and outcome

Administered therapies are displayed in Figure 3. Overall, the treatment regimen of 86.7% of the patients included inhibitors of the RAAS; half of the patients without RAAS inhibitor therapy had received a diagnosis of IgA nephropathy before 1990 (introduction of RAAS inhibitors). In 44.9% of patients, therapy was based exclusively on RAAS inhibitors whereas 46.8% of patients received immunosuppressive therapy in addition to RAAS inhibitor treatment. In nearly all (95.7%) patients, the immunosuppressive regimen included oral and/or intravenous glucocorticoids. The most frequently used non-steroid therapies were cyclophosphamide and azathioprine, followed by mycophenolate mofetil and finally by calcineurin inhibitors.

**Figure 3.**
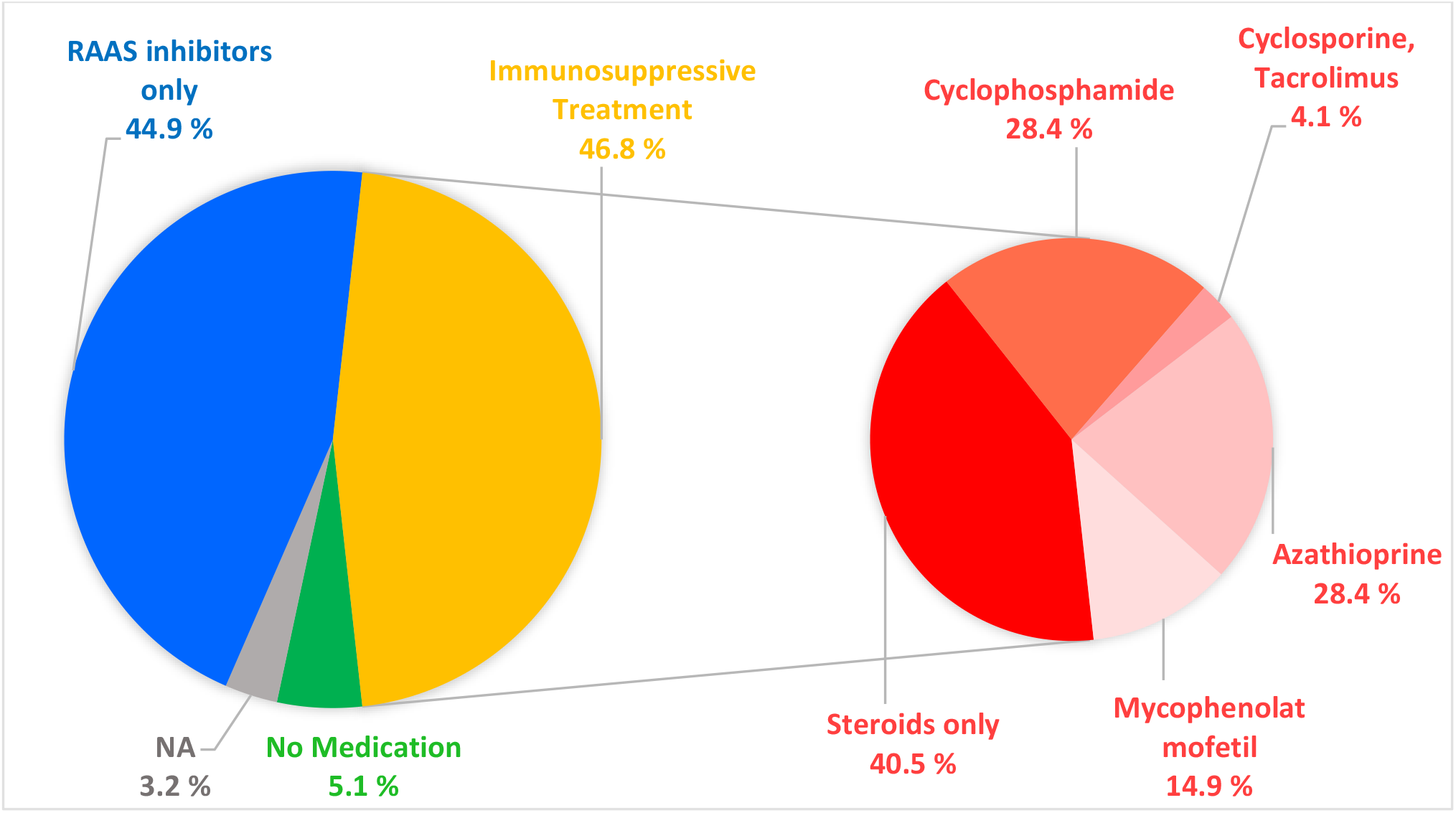
Overall therapy since diagnosis. Values are expressed as percentage of patients receiving specific therapies. RAAS: Renin-Angiotensin-Aldosterone System.

Overall remission rate was 56.3% with 34.1% of all patients fulfilling criteria for complete remission and 22.2% reaching partial remission respectively (Table 2). At the last follow-up, mean serum creatinine was 167.4 µmol/l with a mean eGFR according to CKD-EPI of 54.9 ml/min/1.73 m^2^ and mean proteinuria of 0.8 g/d. Mean systolic and diastolic blood pressure values were 129.5 mmHg and 79.0 mmHg respectively. A total of 43.0 % of patients progressed to ESKD during follow-up, whereas 55.6% of patients developed ESKD within 7 years of follow-up. Among patients not reaching ESKD, 7.9 % had a doubling of serum creatinine value at some point during follow-up. Nearly a third of the patients (31.6 %) received a kidney transplant. One-fifth (18.8 %) of the transplanted patients developed a recurrence of IgA nephropathy within a median follow-up time of 79.7 (10.8 – 457.1) months. Graft rejections occurred in 11.1% of transplanted patients. The mortality rate in the total cohort was 8.9 % within a median follow-up time of 100.0 (0.2-457.1) months.

**Table 2:** Patient outcome.

|  |  |
| --- | --- |
| Blood pressure (mmHg) at last follow-up (n=138) |  |
| - systolic | 129.5 ± 14.9 |
| - diastolic | 79.0 ± 11.5 |
| Serum creatinine (μmol/l) at last follow-up (n=142) | 167.4 ± 144.0 |
| Estimated glomerular filtration rate* (ml/min/1.73 m<sup>2</sup>) at last follow-up | 54.9 ± 28.0 |
| Proteinuria<sup>+</sup> (g/d) at last follow-up (n=144) | 0.8 ± 1.7 |
| Remission after treatment (%) |  |
| - partial remission | 22.2 |
| - complete remission | 34.1 |
| - no response | 43.7 |
| Acute kidney injury at any time point after diagnosis | 12.0 |
| Doubling of serum creatinine (without ESKD) (%) | 7.9 |
| End-stage kidney disease (%) | 43.0 |
| End-stage kidney disease within 7 years (%) | 55.6 |
| Kidney transplantation (%) | 31.6 |
| - graft rejection rate (%) | 11.1 |
| - IgAN recurrence after transplantation (%) | 18.8 |
| Death (%) | 8.9 |
| Median follow-up time (months) | 100.0 (0.2-457.1) |

Values are expressed as means with standard deviation, medians with ranges as indicated or proportions of patients. ESKD: end-stage kidney disease. \*according to Chronic Kidney Disease Epidemiology Collaboration formula. <sup>+</sup> measured from adequately collected 24h-urine collection if available or estimated from urinary protein/creatinine ratio. Acute kidney injury as defined by Kidney Disease Improving Global Outcomes Guidelines 2012.

### Predictors of immunosuppressive therapy use

The comparison between patients treated with and without immunosuppressive therapy showed several differences (Table 3). Immunosuppressive therapy was more frequently prescribed to patients with nephrotic syndrome and higher proteinuria, whereas the presence of extrarenal symptoms did not differ between both groups. In addition, patients with histological findings including presence of crescents were more likely to receive immunosuppressive treatment. Patients treated by immunosuppressive therapy also had by trend higher immunoglobulin staining intensity on kidney biopsy. In contrast, age or kidney function did not significantly affect the decision to apply immunosuppressive therapy. Interestingly, the degree of IFTA was not predictive of immunosuppressive treatment with on the contrary a tendency to higher IFTA scores in patients treated with immunosuppression.

**Table 3:** Group comparison according to use of immunosuppressive therapy.

|  | Immunosuppressive therapy | No immunosuppressive therapy | p value | Corrected p value |
| --- | --- | --- | --- | --- |
|  | (N = 74) | (N = 76) |  |  |
| Age (years) | 52.9 | 53.3 | 0.86 |  |
| Female (%) | 29.7 | 25.0 | 0.52 |  |
| Systolic BP (mmHg) | 138.9 | 138.3 | 0.88 |  |
| Diastolic BP (mmHg) | 85.6 | 82.9 | 0.29 |  |
| eGFR (ml/min/1.73 m<sup>2</sup>) | 49.9 | 60.9 | **0.04** | >0.99 |
| Proteinuria 24h* (g/d) | 3.2 | 1.6 | **<0.001** | **<0.001** |
| Nephrotic Syndrome (%) | 25.0 | 1.5 | **<0.001** | **<0.001** |
| Hematuria (0-4) | 3.2 | 2.6 | **0.03** | >0.99 |
| Extrarenal symptoms (%) | 12.7 | 13.3 | 0.88 |  |
| Crescents (%) | 9.4 | 2.3 | **0.001** | 0.07 |
| Presence of crescents (%) | 54.9 | 21.9 | **<0.001** | **<0.001** |
| Tubular atrophy / Interstitial Fibrosis (%) | 46.0 | 23.9 | **0.007** | 0.48 |
| Tubular atrophy / Interstitial Fibrosis (0-3) | 1.67 | 1.32 | **0.006** | 0.41 |
| IgA staining intensity (0-4) | 2.75 | 2.31 | **0.01** | 0.69 |
| IgG staining intensity (0-4) | 0.81 | 0.45 | **0.005** | 0.35 |
| C3 staining intensity (0-4) | 2.04 | 1.59 | **0.02** | >0.99 |

Values are expressed as means with standard deviation or as proportions of patients. P-values according to student's t-test or Chi square-test as appropriate. Corrected p-values were obtained after applying the Bonferroni method. eGFR: estimated glomerular filtration rate according to Chronic Kidney Disease Epidemiology Collaboration formula. \* measured from adequately collected 24h-urine collection if available or estimated from urinary protein/creatinine ratio.

### Predictors of remission status

The comparison of baseline parameters between the patient subgroups according to remission status is shown in Table 4. At the time of first diagnosis, as expected, macrohematuria, lower systolic blood pressure, higher baseline eGFR, lower proteinuria level and less IFTA were predictors of remission. Nephrotic syndrome did not significantly affect the remission outcome. After correction, only higher baseline eGFR remained as significant predictor of remission.

**Table 4:** Group comparison according to remission status.

|  | Remission | No remission | p value | Corrected p value |
| --- | --- | --- | --- | --- |
|  | (n = 71) | (n = 55) |  |  |
| Age (years) | 53.4 | 55.0 | 0.56 |  |
| Female (%) | 29.6 | 20.0 | 0.22 |  |
| Systolic BP (mmHg) | 135.3 | 145.5 | **0.02** | >0.99 |
| Diastolic BP (mmHg) | 82.3 | 87.5 | 0.07 |  |
| eGFR (ml/min/1.73 m<sup>2</sup>) | 60.6 | 40.3 | **<0.001** | **0.02** |
| Proteinuria 24h* (g/d) | 2.2 | 3.2 | **0.02** | >0.99 |
| Nephrotic Syndrome (%) | 11.1 | 19.5 | 0.23 |  |
| Macrohematuria (%) | 39.1 | 18.5 | **0.01** | 0.9 |
| Crescents (% glomeruli) | 5.9 | 6.7 | 0.76 |  |
| Presence of crescents (%) | 42.3 | 35.3 | 0.44 |  |
| Tubular atrophy / Interstitial Fibrosis (%) | 29.9 | 48.9 | **0.04** | >0.99 |
| IgA staining intensity (0-4) | 2.55 | 2.59 | 0.82 |  |
| IgG staining intensity (0-4) | 0.77 | 0.51 | 0.08 |  |
| C3 staining intensity (0-4) | 1.81 | 2.19 | 0.12 |  |

Values are expressed as means with standard deviation or as proportions of patients. P-values according to student's t-test or Chi square test as appropriate. Corrected p-values were obtained after applying the Bonferroni method. eGFR: estimated glomerular filtration rate according to Chronic Kidney Disease Epidemiology Collaboration formula. \* measured from adequately collected 24h-urine collection if available or estimated from urinary protein/creatinine ratio.

### Predictors of progression to end-stage kidney disease

Patients who progressed to ESKD were on average older and had higher blood pressure, lower eGFR and higher proteinuria level at baseline as compared to non-progressors (Table 5). In addition, patients reaching ESKD more frequently exhibited nephrotic syndrome at the time of diagnosis, while clinical remission had occurred less frequently. Overall, 13.3% of patients with proteinuria <1g/d at baseline developed ESKD in this cohort. Histologically, more extensive IFTA and crescents were present at baseline in progressors. After correction, lower baseline eGFR, higher proteinuria and higher extent of IFTA remained predictors of progression to ESKD.

**Table 5:** Group comparison according to progression status.

|  | ESKD reached<br>(N = 68) | No ESKD reached<br>(N = 89) | p value | Corrected<br>p value |
| --- | --- | --- | --- | --- |
| Age (years) | 57.2 | 50.5 | **0.004** | 0.28 |
| Female (%) | 22.1 | 29.2 | 0.31 |  |
| Nephrotic syndrome (%) | 20.9 | 7.5 | **0.03** | >0.99 |
| Systolic BP (mmHg) | 146.0 | 134.5 | **0.005** | 0.35 |
| Diastolic BP (mmHg) | 89.0 | 81.6 | **0.006** | 0.41 |
| eGFR (ml/min/1.73 m2) | 41.1 | 64.2 | **<0.001** | **<0.001** |
| Proteinuria 24 h (g/d) | 3.4 | 1.8 | **<0.001** | **<0.001** |
| Crescents (% of total) | 8.8 | 3.8 | **0.03** | >0.99 |
| Presence of crescents (%) | 16.6 | 29.2 | 0.23 |  |
| Tubular atrophy / Interstitial Fibrosis (%) | 53.7 | 20.2 | **<0.001** | **<0.001** |
| IgA staining intensity (0-4+) | 2.60 | 2.45 | 0.38 |  |
| IgG staining intensity (0-4+) | 0.72 | 0.54 | 0.16 |  |
| C3 staining intensity (0-4+) | 2.00 | 1.72 | 0.25 |  |
| Overall remission (%) | 34.0 | 78.1 | **<0.001** | **<0.001** |
| Partial remission (%) | 22.6 | 21.9 | 0.92 |  |

Values are expressed as means with standard deviation or as proportions of patients. P-values according to student's t-test. Corrected p-values were obtained after applying the Bonferroni method. EGFR: estimated glomerular filtration rate according to Chronic Kidney Disease Epidemiology Collaboration formula. \* measured from adequately collected 24h-urine collection if available or estimated from urinary protein/creatinine ratio.

## Discussion

The goal of this study was to perform a detailed review of all patients with IgA nephropathy treated at our Swiss tertiary center focusing on disease characteristics, treatment practices and outcome as well as to evaluate potential predictors of therapeutic choices and patient outcome. The main findings were the following: i) a large spectrum of age-dependent clinical manifestations, ii) progression to ESKD in a comparatively high proportion of patients, and iii) use of immunosuppressive therapy, particularly non-steroid immunosuppressive therapy, in an important part of patients.

First, in this cohort of mainly Caucasian patients exhibiting an expected sex distribution, a wide range of clinical presentations was noted including isolated urine abnormalities, nephrotic syndrome, chronic kidney function impairment and AKI as previously reported [5]. However, chronic kidney function impairment was by far the most common clinical presentation. This may be due to the older patient age and the advanced stage of disease at diagnosis (i.e. higher proportion of patients with moderate to severe IFTA) as compared to other cohorts [25]. Indeed, in this cohort, disease presentation varied considerably according to age. As expected, presentation with macrohematuria as sole disease manifestation was more frequent in younger adults decreasing in frequency with age. In addition, in this cohort, younger adults more often presented with asymptomatic urine abnormalities. Isolated microhematuria was not noted since it usually does not represent an indication for kidney biopsy in Switzerland. Interestingly, in this cohort, the frequency of extrarenal symptoms pointing to a diagnosis of Henoch-Schönlein purpura was not significantly different among different age groups whereas a usually higher frequency is described in younger patients [26]. However, this might be explained by the exclusion of children from this cohort and more frequent secondary etiologies in adults including drugs, infection and cancer becoming more common with age. Lastly, in this cohort, nephrotic syndrome classically described as an uncommon clinical presentation in IgA nephropathy, was noted at baseline in roughly 10% of patients and more frequently so in women. This is in contrast to data from Chinese patients with IgA nephropathy indicating more severe clinical presentation including proteinuria level in male patients [27]. The reasons for this discrepancy are not clear. Although spontaneous remissions have been described in the setting of nephrotic syndrome in IgA nephropathy and to be more frequent in female patients according to a Korean study, in our cohort, the presence of nephrotic syndrome was more often observed among patients treated with immunosuppressive therapy [28].

Second, a high rate of progression was noted in our cohort, with over half of the patients reaching ESKD within 7 years of follow-up, 43% of the patients during a mean follow-up time of 8.3 years. In contrast, 18.8% of patients in the European VALIGA cohort progressed to ESKD during a median follow-up time of 7 years, 12.6% of patients in the German CKD cohort over 6.5 years, while 34% in a Norwegian cohort reached a combined end-point of ESKD or death within 8 years [29-31]. Similarly, lower rates of progression are reported among non-Caucasian patient cohorts [32]. This finding might be explained by the rather advanced presentation at baseline in our cohort including lower eGFR, higher proteinuria, higher blood pressure and greater extent of chronic interstitial lesions on kidney biopsy [25, 30-32]. Indeed, in our cohort, lower eGFR and higher proteinuria at presentation were confirmed as predictors of progression to ESKD. In line with this hypothesis, a similarly poor outcome has been reported recently in a UK cohort of patients with comparably severe clinical presentation [33]. In addition, the comparatively older age of patients in our cohort might have contributed to worse outcomes [25, 30, 32]. Finally, the inclusion of patients treated at a tertiary clinical center might have led to selection of more severe cases. Indeed, the clinical presentation with macroscopic hematuria in our cohort was much less frequent than described previously [5]. It is further important to note that 13.3% of patients presenting with proteinuria <1g/d in this cohort progressed to ESKD during follow-up extending recent findings from the UK RaDaR cohort correlating time-averaged proteinuria to development of ESKD [33].

Third, almost half of the included patients had received immunosuppressive treatment for IgA nephropathy. In comparison, 19-25% of patients had received immunosuppressive treatment in the German CKD cohort and a Chinese glomerulonephritis registry [31, 32]. Although this proportion might therefore appear high, a comparable proportion of patients had received immunosuppressive therapy in the VALIGA cohort. This might be explained by the major contribution of Italian centers to the last-mentioned cohort with traditional comparatively higher use of immunosuppressive therapy in the treatment of IgA nephropathy [18, 19, 25]. Furthermore, however, more than half of the patients in our cohort had received a non-steroid-based immunosuppressive regimen, which corresponds to a clearly higher proportion of patients than in the VALIGA cohort [25]. Among the employed therapies, cyclophosphamide and azathioprine were the most common. As expected, higher proteinuria represented a main predictive factor for the decision to use immunosuppressive therapy in these patients. Additionally, 28.9% of the patients with proteinuria <1g/d at presentation had received immunosuppression for the treatment of IgA nephropathy. However, the presence of extrarenal symptoms did not differ between the groups treated with or without immunosuppression. It can be speculated that the more severe presentation at baseline in our cohort including higher proteinuria levels might have favored the use of immunosuppressive therapies. In addition, as referral center, therapy-refractory disease or patients pre-treated with non-immunosuppressive and steroid-based regimens may have been disproportionally represented. The frequent use of azathioprine might also be explained by the participation in an international randomized-controlled trial investigating the effect of azathioprine in the treatment of IgA nephropathy [34]. The coverage by RAAS inhibitor treatment in our cohort was large and comparable to other cohorts taking into account the small proportion of patients treated before the advent of this drug class.

Interestingly, the degree of chronic changes on the initial kidney biopsy (i.e. IFTA) did not predict utilization of immunosuppressive therapy in this cohort. In contrast, an opposite trend was found with patients receiving an immunosuppressive treatment having by trend higher percentage of IFTA at baseline. The reasons for this finding are unclear. The degree of IFTA did not represent an exclusion criterion in the randomized-controlled trial evaluating azathioprine mentioned above [34]. However, no difference was observed regarding IFTA severity in patients treated with azathioprine and the rest of the cohort. A limiting factor here might be the sample size, which may not have been large enough to analyze this hypothesis. Other factors identified as predictors of the use of immunosuppression in patients with IgA nephropathy in our study such as proteinuria level, nephrotic syndrome and the presence of crescents were expected and mirror the findings of a recent survey [21].

Limitations of this analysis have to be considered. Firstly, the retrospective design precludes reconstruction of therapy choices although significant differences among treatment groups in this cohort point to factors influencing therapeutic decisions. Secondly, analysis from a single center cohort in a tertiary center may have led to selection bias and reduced generalizability. Thirdly and similarly, in this cohort of mainly Caucasian patients, results have to be interpreted within this context. Fourthly, in this analysis, merely univariate analysis of predictors has been performed, however controlling for multiple comparisons using a conservative method. Fifthly, our patient cohort is of limited sample size. However, despite this, significant predictors of therapy and outcome were identified.

In conclusion, at the dawn of a new therapeutic era for patients with IgA nephropathy [35], this retrospective cohort analysis from 1980 – 2017 gives detailed insight into clinical and histological characteristics and outcome of patients with IgA nephropathy from a Swiss tertiary center as well as treatment practices and potential predictors of outcome and therapy.

## Acknowledgements

Ethical approval was provided by the cantonal ethics committee of the canton of Bern, Switzerland.

## Funding

This work was funded by the University clinic for Nephrology and Hypertension, Inselspital University Hospital Bern, Bern, Switzerland.

## Data availability statement

The data underlying this article will be shared on reasonable request to the corresponding author.

## Authors’ contributions

L.-Y.M.: Research idea, study design, supervision, data analysis, interpretation of data, writing, review and editing. D.T.T.: Data collection, data analysis, creation of figures and tables, writing (first draft), review and editing. B.V.: Review, validation. All authors approved the final manuscript.

## Notes

### Competing Interest Statement

The authors have declared no competing interest.

### Author Declarations

Ethics committee of the Canton of Bern gave ethical approval for this work.

